# Confidential Screening for and Management of Intimate Partner Violence Risk using a Commercial Electronic Health Records System

**DOI:** 10.1101/2023.02.11.23284286

**Authors:** Leslie A Lenert, Vanessa Diaz, Kit Simpson, Christine Hahn, Michael Aiken, Elizabeth Swazt, Luke Sox, Ekaterina Pekar, Naomi Ennis, Alyssa Rheingold

## Abstract

Intimate partner violence is, unfortunately, a common problem (25% of women) for which screening in primary care is a recommended service. In this paper, we describe modifications to a commercial EHR system (Epic) designed to support confidential screening for and management of IPV in primary care settings. Modifications include the use of an exam room computer as a kiosk for patient-generated health data entry, storage of data in a hidden location, the use of rule-based alerting methods to direct providers to access data, and electronic form-based tools for case management and documentation. While preserving privacy, this approach also allows access by provider type and authorized setting, including use for population health management. The approach was tested in a pilot study and found to be feasible, to have good compliance for provider screening (65%) and is being evaluated in a stepped-wedge trial in other primary care clinics across a large academic health system.

## Introduction

Intimate Partner Violence (IPV) is a frequently occurring event (25% lifetime prevalence) for which screening in primary care is the United States Preventative Service Task Force recommended service [1]. Despite its prevalence, screening for IPV is conducted much less frequently in primary care than screening for other medical conditions such as depression [2] This is because there are significant barriers to screening. Two convolved barriers, addressed in this paper, are a lack of privacy in the electronic health record for issues surrounding IPV and a lack of support for assessment, management and referral for subsequent care. We describe herein, modifications to a commercial electronic records system (Epic) that enable confidential screening, assessment, management and referral for IPV.

While IPV poses risks, its victims also fear stigmatization. This is a significant barrier to disclosure: for example, some women in emergency department settings with obvious evidence of injuries, chose not to disclose partner abuse because of privacy and safety concerns [3]. Concerns about confidentiality are a recognized barrier to disclosure in screening [4]. The usual approaches to screen for medical issues, such as the verbal administration of questions by providers may not be as effective in this area. For example, if providers ask IPV-related screening questions in a routine or uncaring way, it can put off patients and discourage honest responses [2]. An alternative approach to screening is to use self-administered computer questionnaires, which is an effective but under-utilized method [5]. However, these methods have an important practical problem–it is difficult to offer IPV victims sufficient privacy during clinical visits for forthright responses. Potential perpetrators could be in the exam room during screening and tablets in the waiting room might not afford sufficient privacy. Screening by sending questionnaires to a patient’s phone might offer sufficient privacy; however, there are a growing number of “stalker” apps that could be installed by a perpetrator on the patient’s phone to monitor a patient’s activities online [6].

After screening, providers need to assess the risk of future violence and take actions to mitigate which need to be documented. This can pose additional threats of loss of confidentiality. Moreover, few primary care providers have adequate training in the assessment and management of IPV-related risk. EHR-based decision support tools may help providers feel more comfortable with screening, assessment and management tasks [4,7], and thus encourage compliance with screening guidelines. However, this creates further issues with data privacy. If the data generated by IPV management decision support applications generate electronic health record (EHR) documentation that is easily viewed in the record or shared in medical record releases, this could inhibit disclosures [2,8] or pose additional risks to the victims. An IPV-related diagnosis might be inadvertently disclosed to a perpetrator in a number of ways. For example, IPV diagnoses could appear in printed post-visit summaries for patients, in queries of a 21st Century Cures Act interfaces to the EHR by apps, in medical records releases, and in codes reported to support claims data.

In EHR systems, one approach to preserve privacy for stigmatizing conditions is the segmentation of data from the general medical record. In specialty care, management of confidential conditions such as drug abuse can occur in secure “departments” matched to organization units, that prevent record viewing by other providers. In some circumstances, specific providers not in the organizational unit can have what is called “break the glass” capability in the Epic EHR for medical emergencies, which allows a provider to view segmented parts of the record but also creates a record of the event in a log file and/or a notification event sent to a data security officer.

In primary care settings, where patient care typically involves care of multiple problems at the same time, maintaining differential patient privacy while screening for and managing a condition *confidentially* is difficult to implement. Segmentation of specific data elements has not been widely implemented in commercial EHR systems. For example, while drug abuse issues in a drug addiction treatment clinic might be protected by “break the glass”-like functionality, in primary care settings, entry of a problem list diagnosis of opioid abuse (F11.1) would make it available broadly across a healthcare enterprise and result in its inclusion in summary documents and disclosed medical record requests. Questionnaire responses in patients who experience physical or mental abuse in a relationship, or even the act of screening for such issues, could result in the introduction of specific codes for such into the medical record such as Z91.41 (personal history of adult physical and sexual abuse) into parts of the medical record, especially if physicians document the screening or counseling in their notes. Even so, because the essence of primary care is the longitudinal follow-up of patients over time, it is critical to somehow notify a provider of a prior history of positive screening results for IPV and encourage periodic reassessment.

While screening and counseling for IPV is a billable activity, the desire of providers to keep discussions as private as possible may result in the provider not using an IPV-related billing code and losing potential credit for time spent counseling in provider productivity calculations. An approach to document time spent in confidential activities that a health system might *choose not to bill for* because of privacy concerns may be important to incentivize providers’ screening for IPV.

In this paper, we describe modifications to a commercial EHR, Epic, designed to allow its use to screen for and care for a confidential condition while maintaining differential data privacy in a primary care setting. The approach is pragmatic but comprehensive, screening patients privately using self-reported questionnaires and preventing accidental disclosures in visit summaries, billing records, and releases of medical records but also supporting providers in assessments and allowing providers to track at-risk patients, assess and document individual risk, document time spent on IPV care and provide follow up initial care for IPV positive screening patients. The approach uses pop-up-style alerts to notify providers of IPV screening requirements and of prior data while storing results in a segmented part of the record.

## Methods

Working with key stakeholders (i.e., providers, patients, IPV survivors, and IPV experts) we adapted an existing program for IPV screening with both paper-based and computer-based components used within Northern California Kaiser Permanente Health System [9] to be fully implemented within the EHR *in a privacy-preserving manner*. The implementation included the creation of a registry of targeted patients (women 18 to 50 years of age, who are the highest risk group) being seen in primary care clinics on an annual basis. The resulting workflow was complex and included population health, patient data input, and provider input components as shown in Figure 1. The EHR’s registry monitored patients meeting inclusion criteria and triggered in-menu alert notifications to medical assistants reminding them to screen a patient for IPV on an annual basis using a non-interruptive alert (Figure 2).

**Figure 1.**
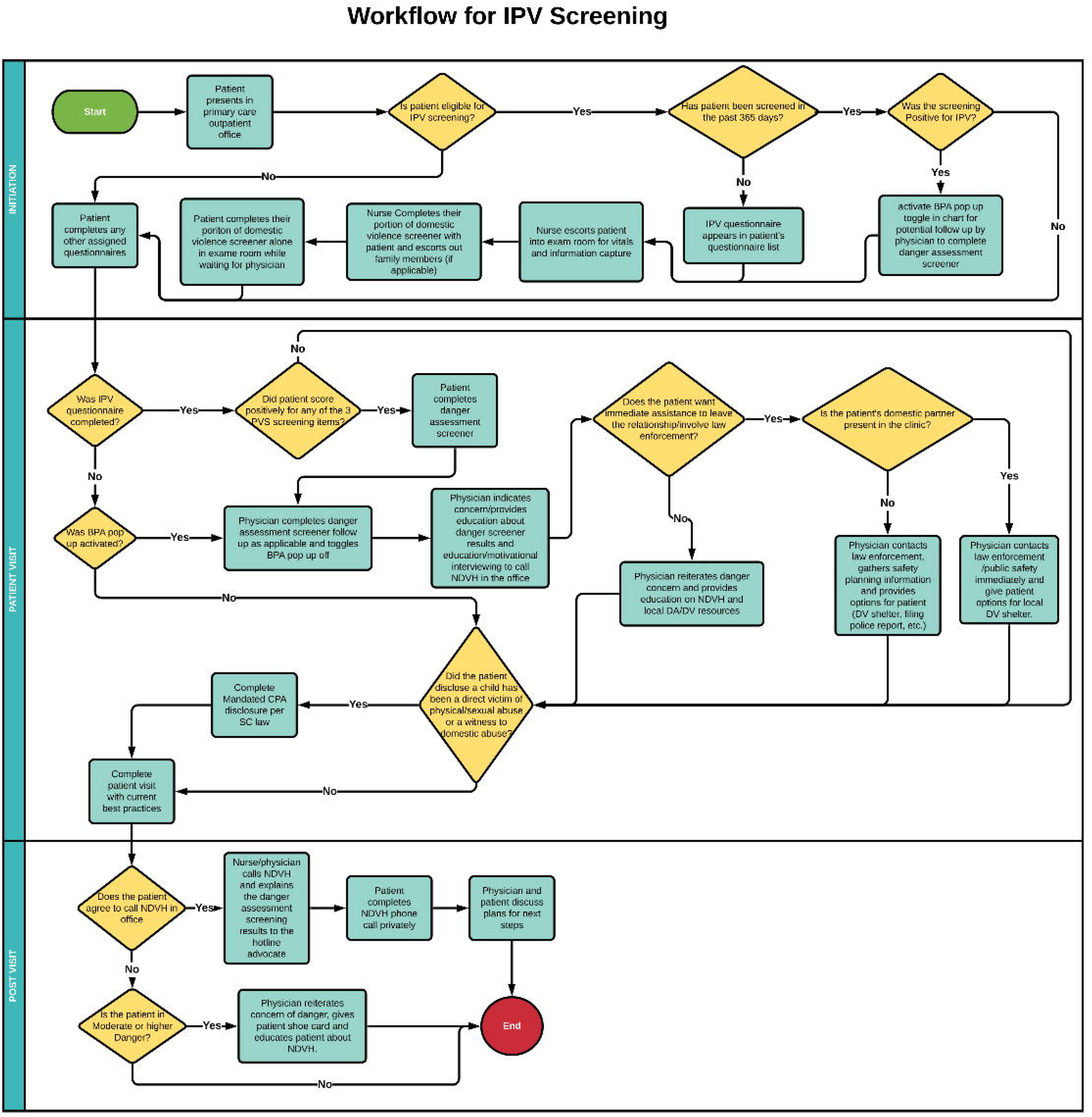
Workflow for IPV screening, risk assessment and management.

**Figure 2.**
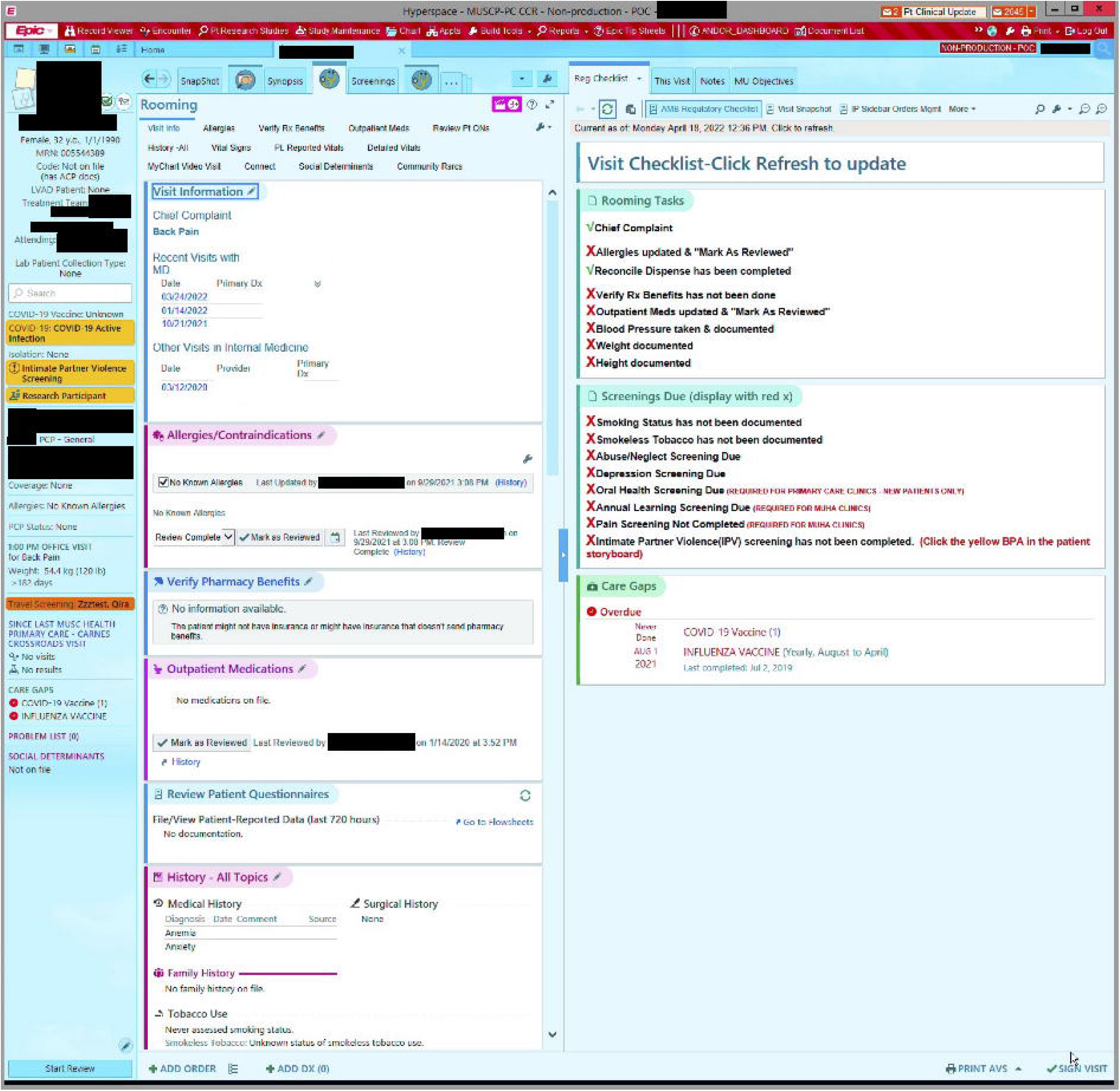
Summary of Screening questionnaire responses displayed. Used with Permission of Epic Systems.

Responding to the alert converted the exam room computer to a kiosk-like mode for a patient to self-administer a questionnaire. The medical assistant assured that the patient was alone, by removing another adult-age family from the room, to allow private responses to the 3-item initial screening questionnaire [10]. If positive, additional questionnaires were administered to assess risk levels [11] (figure 3).

**Figure 3.**
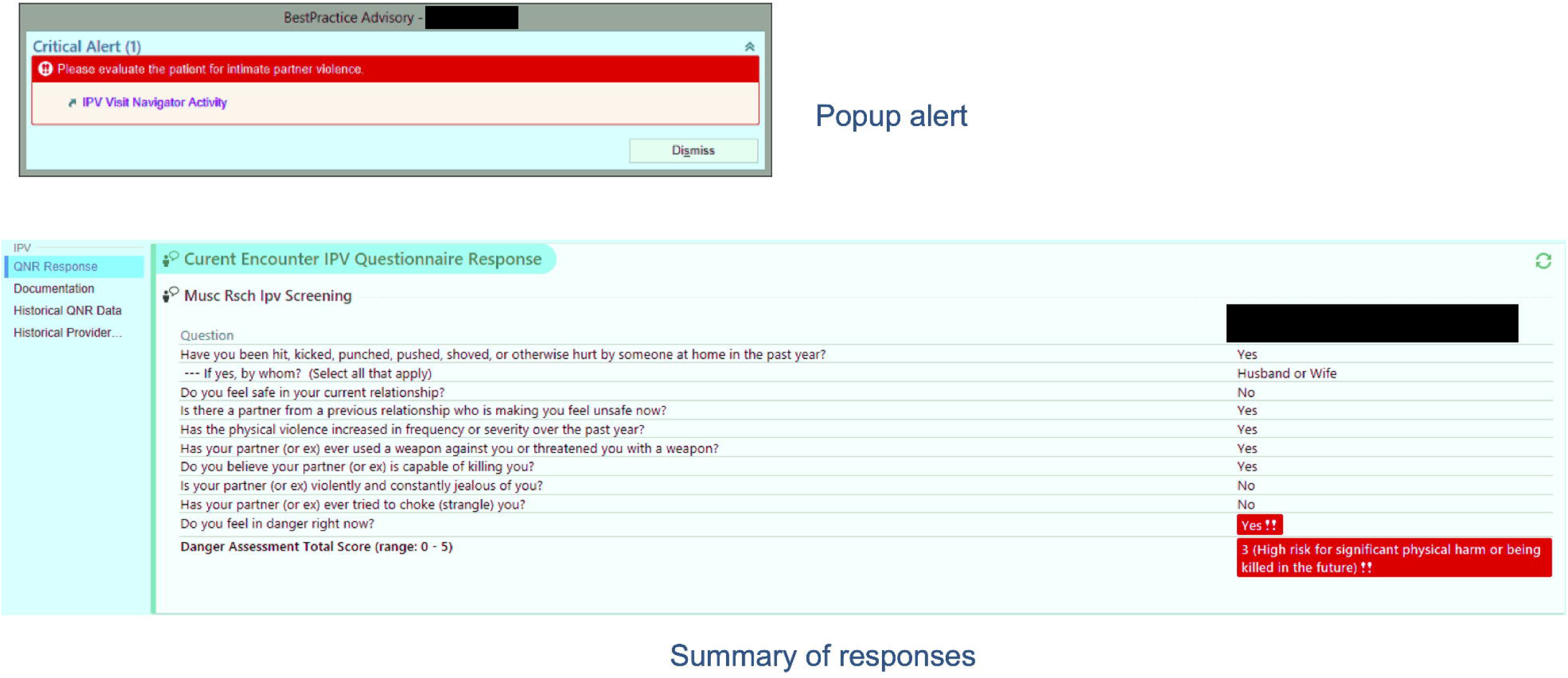
Provider tool for the assessment of risk for patients screening positive for IPV risk, documentation of physical examination findings, and for referral for additional counseling or care. Used with Permission of Epic Systems.

Completion of the questionnaires resulted in a lock screen on the exam room computer to await the provider’s login for their portion of the visit. If the patient has screened positive for IPV, a second, blocking style popup alert notified the provider and took the provider to an electronic form that reviewed the patient’s responses (figure 4), and helped evaluation of risk levels, additional recommended follow-up questions with example scripted responses, documentation of counseling on the patient, and of time spent on this issue during the visit (Figure 5).

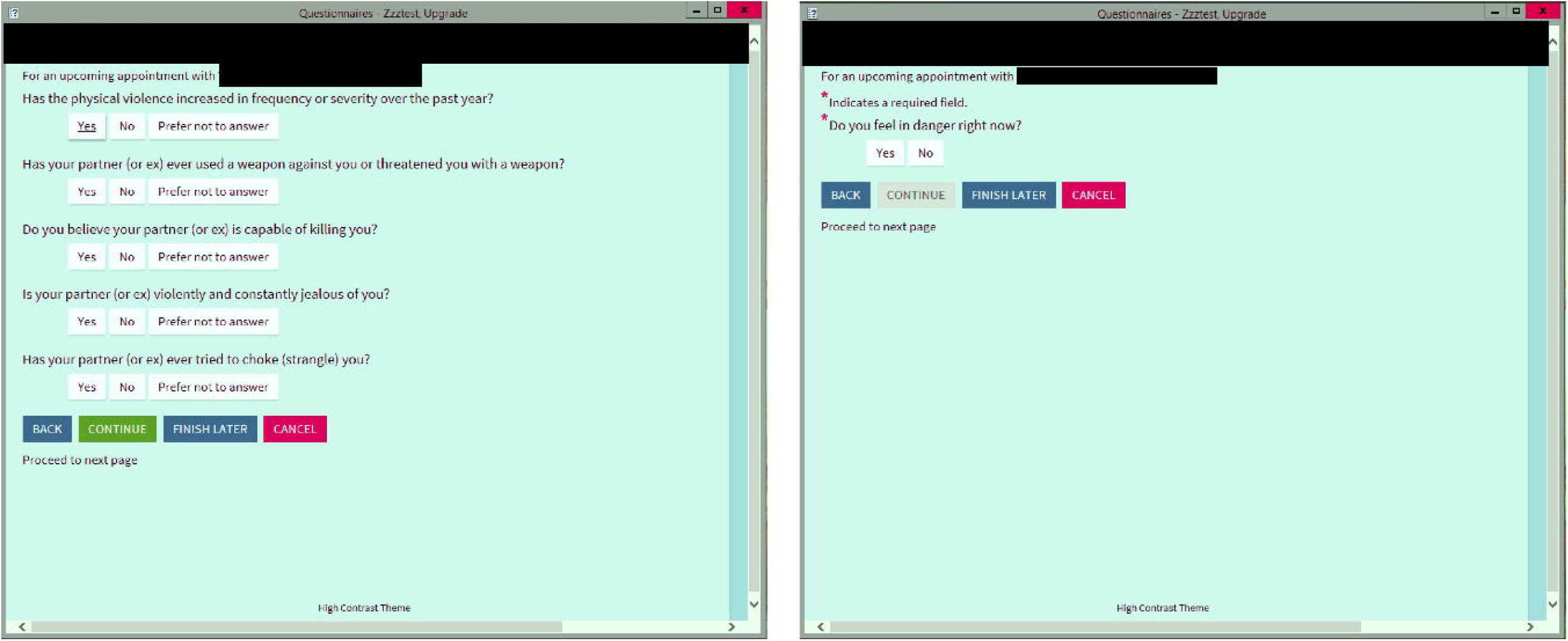

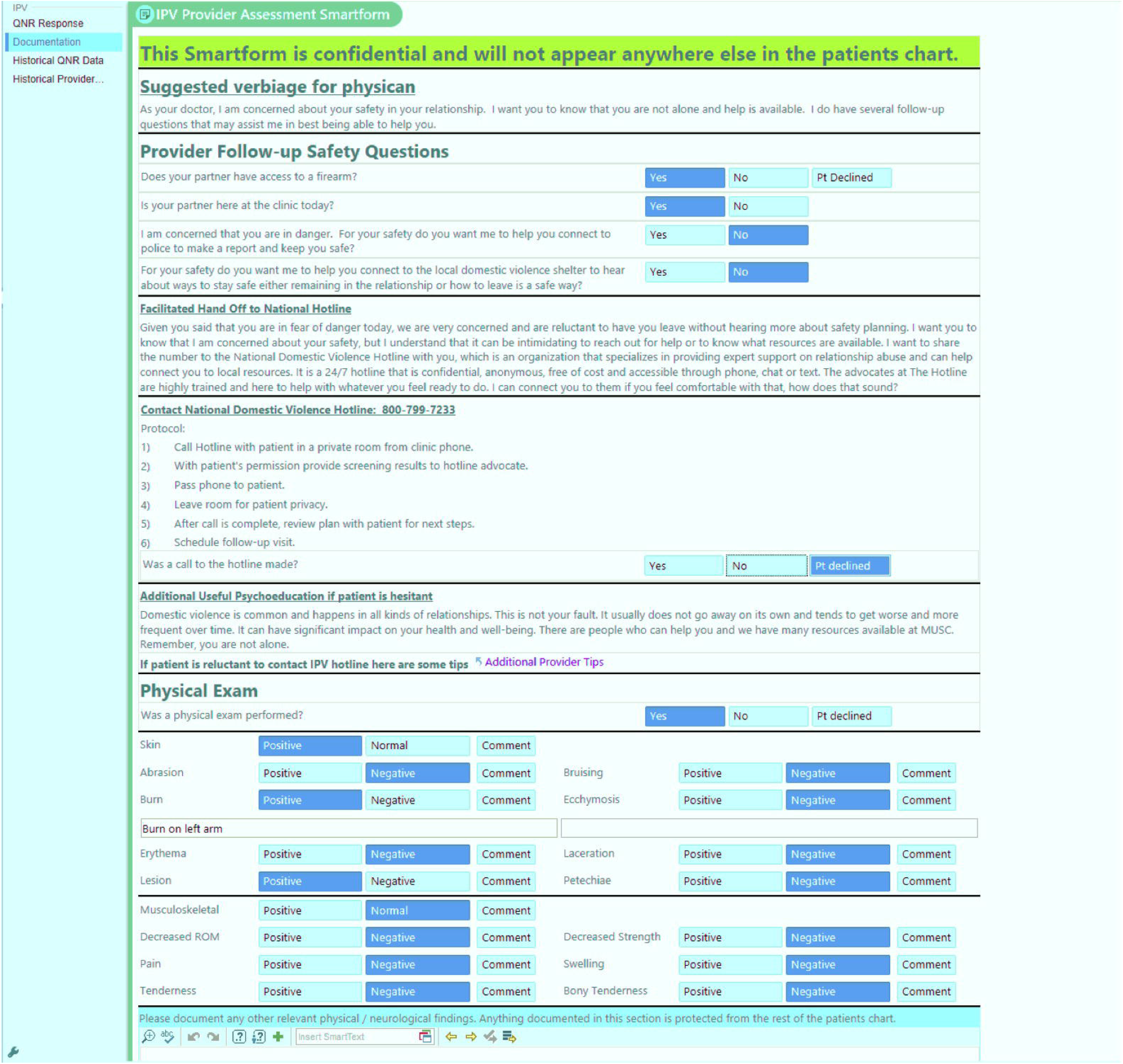

The results were stored in an electronic flow sheet segmented from other visit records. On return visits, patients with positive prior screenings are rescreened and providers were directed to the flow sheet to review prior data by a different blocking-style popup alert. Providers then completed reassessments of risk, physical exam findings, and document follow-up recommendations.

Referral for post-visit care was managed in two ways. In settings of immediate risk to the patient, there was a protocol for immediate outreach to hospital security and a nurse specialist providing IPV support to the Emergency Department. For lower-risk cases, patients were referred to a national hotline for counseling and support for IPV victims. In cases where the abuse of the victim was observed by children, the provider was reminded in the documentation tool to immediately refer the case to state child protective services, as required by law, and contact information was given.

After pilot testing using simulated patients in a controlled setting, and further refinement of workflows for interactive assessment of IPV risks by a provider, we implemented our screening program in a primary care clinic. The staff was trained in the use of the IPV screening system using in-person and online tools. Educational materials included tip sheets on intimate partner violence and how to implement screening procedures, video demonstrations of EHR use, as well as videos demonstrating suggested scripting for interactions with patients. Clinic compliance with screening was monitored weekly with discussions with clinic leadership and retraining provided. No other incentives for the adoption of the screening program were offered. The study was implemented as a quality improvement project and deemed exempt from human subjects review on that basis by a University institutional review board.

## Results

The pilot phase began on March 4^th^ of 2020. Unfortunately, COVID-19 social distancing policies at MUSC resulted in the closure of almost all in-person operations in the pilot clinic (and as a result, cessation of screening) on March 19th. During this two-business-week period, there were 57 eligible patients seen. Of these, 38 had screening initiated and 37 patients finished screening 65% compliance. Two patients screened positive and were further evaluated. Both patients were fully assessed by the providers in response to the pop-up alert. In both cases, the provider documented that it required between 0-15 minutes of additional time to complete counseling. Data collection during in-person visits continued in the pilot clinic until the end of the first quarter, as patients continued to be seen intermittently.

After the reopening of in-person clinic operations, we began a stepped wedge design [12] rollout across 20 primary care clinics in 2021. Table 1 shows interim results over the first two iterations of the stepped design. Uptake across clinics after the restoration of operations was lower than in the pilot phase, except for one clinic, and clinic adoption was widely disparate despite the efforts of the investigators to encourage adherence. Lower uptake was seen in the context of reduced clinic staffing due to the ongoing Covid-19 emergency. One clinic administrator thought the questionnaire administration process was too disruptive to clinical workflows to adopt and refused to participate.

**Table 1.**
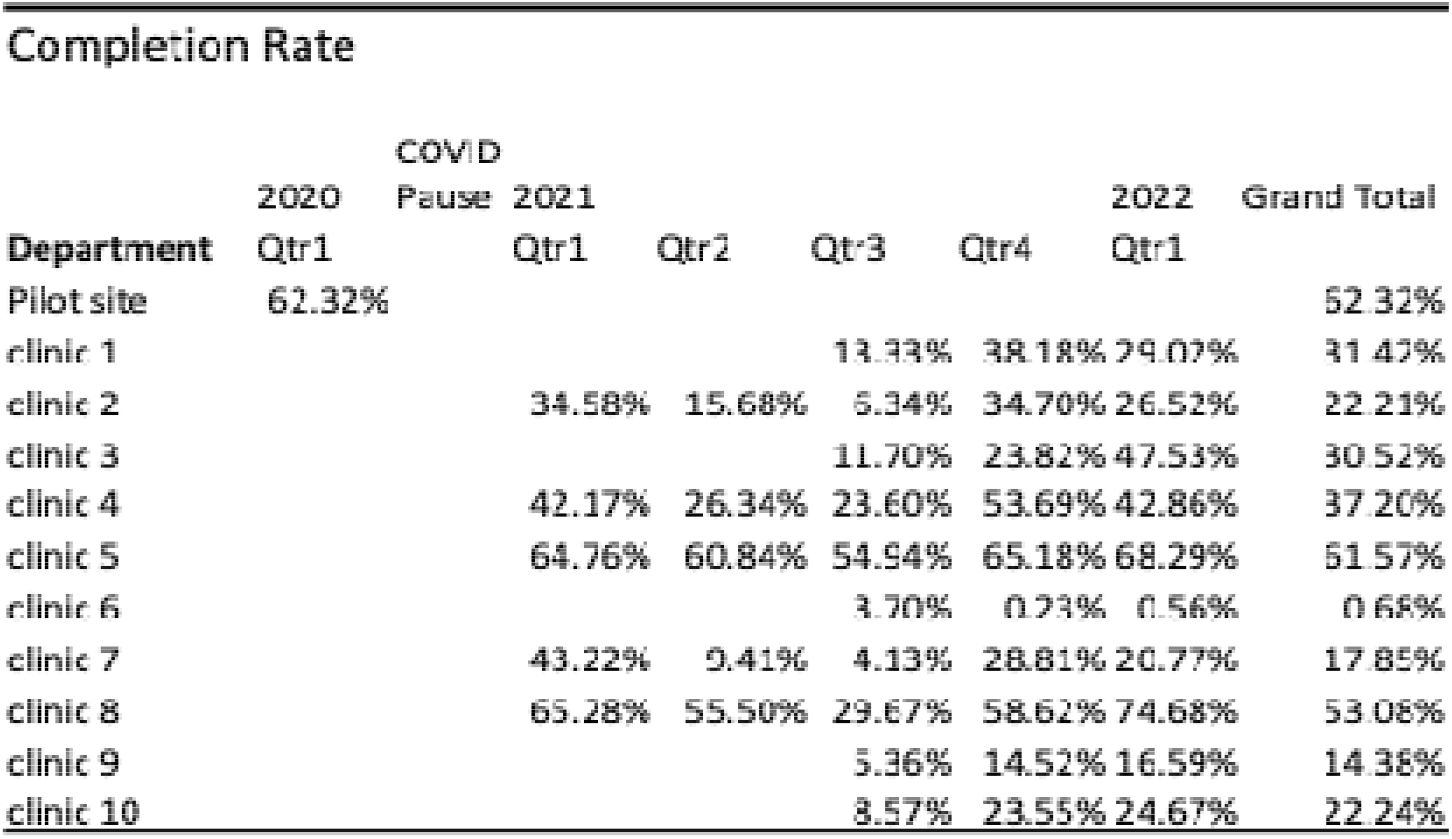
Interim analysis of clinic compliance.

## Discussion

This paper describes a pragmatic approach to screening for confidential conditions and segmentation of subsequent data in the clinical record with preservation of the ability to use that data in subsequent clinical care and population health management. The approach uses alert triggers linked to pop-up reminders of critical data in the chart and the need for assessment or follow-up. This approach takes advantage of the greater flexibility provided by decision support tools in the EHR to limit the presentation of data to providers in specific circumstances, as specified in the rule. This workaround allows the designation and the maintenance of differential privacy in the record within a visit. Visualization of data in the record uses a combination of an EHR Flow Sheet data structure as a time-oriented variable-sized data container, in combination with Epic specific tool, a “SmartForm” software object that retrieves the data and presents in a manageable way for use in primary care practices, as well as providing support for assessment.

The use of pop-up alerts allows both clinic-specific and personnel-specific constraints on access. It also can remind providers to view relevant confidential data when caring for persons in a different context. For example, a pop-up alert could notify an emergency department provider or a radiologist to view IPV risk data when reviewing evaluating a patient. Tracking of access to confidential data for enforcement of privacy restrictions, which is the critical feature of “break-the-glass” functionality, is not lost by using a popup alert-driven approach. Providers can be tracked by mechanisms that record responses to pop-up alerts in the EHR for quality improvement purposes. However, one limitation of the approach is that tracking of access by browsing of electronic flow sheet data in the EHR is not possible.

Explicitly modeling security at a data element level is not a feature of current commercial EHRs. A trial in an experimental version of the Regenstrief Institute’s CareView system (now replaced by a commercial system), demonstrated the feasibility of patients designating certain sections for restricted access by providers, and the potential acceptance of providers of this approach [13,14].

Recent changes in privacy protections for women’s healthcare have come about with the *Dobbs vs. Jackson Women’s Health Org ruling*, which severely limits the rights of privacy established in Roe vs. Wade [15]. These changes highlight the need for differential privacy in health records for women’s health issues beyond IPV. While this application was designed to enhance privacy for better reporting and patients’ safety, there are now legal issues that should prompt EHR developers and policymakers to take a careful look at differential privacy in the record. As currently mandated by the 21st Century Cures Act and implemented within certified EHR systems, given a request for a medical record using electronic means, a provider is required to transfer a complete electronic record of care [16]. This could be problematic for both patients and their healthcare providers if the provision of that care is unlawful in the state of the requestor. Clayton and colleagues have proposed a variety of ad hoc solutions for responding to this challenge [17]. Approaches such as shadow records and clinic-specific paper charts may prevent disclosures but also would lock information away to prevent its positive uses, such as population health practices for subsequent management or alerts to other providers. Segmentation of data in the EHR record with infrastructure to prevent transmission unless authorized by the patient and provider could address many issues related to inadvertent disclosure. Our work shows how this could be implemented today in an existing commercial EHR.

## Limitations

Segmentation of data in custom flow sheets in an EHR does create new issues as to what constitutes the official medical record in an EHR system. Pop-up alerts may have to be implemented for the specific care settings to inform providers that need to be aware of screening results. These interruptive alerts may be missed by providers, as compliance with this type of alert is not consistent. On the other hand, a specific alert for positive data on IPV risk may be very helpful in certain settings to direct a provider’s attention to the issue of a non-accidental cause of an injury, for example, in the emergency department. This approach does not preclude a provider from documenting IPV in the record if he or she feels it is appropriate to do so.

The rate of adoption of the highly confidential screening workflow was initially good (67%) but fell in a post-COVID-19-peak setting to less than optimal in rollouts to additional clinics. The screening was voluntary and as a result, charts could be closed without doing the screening. There was consideration given to including the IPV screening component as a required operation similar to some other health screening areas but based on feedback from stakeholders, we opted to keep screening voluntary. A greater ability to customize patient-generated data collection to clinic workflows could improve adherence rates. The current approach tracks provider time for counseling but does not provide further incentives. If incentive programs for clinic and provider staff for compliance were added to this implementation, results could further improve.

## Conclusions

Practical approaches to segment EHR data to enhance patients’ privacy are critically important to develop. This study demonstrates the feasibility of secure confidential screening of patients for intimate partner violence in primary care settings, and secure confidential management of data on IPV risk in the EHR. Results from an ongoing study will assess the impact of the approach and the feasibility of scaling the approach across an enterprise.

## Data Availability

All data produced in the present study are available upon reasonable request to the authors

## Contributorship

LL and AR conceived of the study, obtained funding, supervised implementation, and contributed to the authorship of the manuscript. VD and KS contributed to the design of the study and to the authorship of the manuscript. VD had a lead role in the implementation of the study. CH, MA, ES, LS, EK, and NS contributed to the design and implementation of the study and to editing of the manuscript.

## Acknowledgments

We are grateful to BMIC and clinical team members for their support that made the study possible.

## Software availability

The software is available to other Epic customer institutions on request to.

## Data availability

Data are available on written request to.

